# Low Dimensional Chaotic Attractors in Daily Hospital Occupancy from COVID-19 in the USA and Canada

**DOI:** 10.1101/2022.12.04.22283069

**Authors:** Carlos Pedro Gonçalves

## Abstract

Epidemiological application of chaos theory methods have uncovered the existence of chaotic markers in SARS-CoV-2’s epidemiological data, including low dimensional attractors with positive Lyapunov exponents, and evidence markers of a dynamics that is close to the onset of chaos for different regions. We expand on these previous works, performing a comparative study of United States of America (USA) and Canada’s COVID-19 daily hospital occupancy cases, applying a combination of chaos theory, machine learning and topological data analysis methods. Both countries show markers of low dimensional chaos for the COVID-19 hospitalization data, with a high predictability for adaptive artificial intelligence systems exploiting the recurrence structure of these attractors, with more than 95% *R*^2^ scores for up to 42 days ahead prediction. The evidence is favorable to the USA’s hospitalizations being closer to the onset of chaos and more predictable than Canada, the reasons for this higher predictability are accounted for by using topological data analysis methods.

## 1. Introduction

Epidemiological applications of chaos theory methods to the SARS-CoV-2/COVID-19 pandemic [1-5] have uncovered evidence of chaotic markers in the pandemics’ dynamics. In [5] we found evidence of stochastic chaos with emergent low dimensional attractors for the COVID-19 regional data at the level of the number of new positive cases per million and the number of new deaths per million, in particular, the type of dynamics identified was a power law chaos dynamics also called color chaos [5-7], characterized by power law signatures in the frequency spectrum, this dynamics occurred for Asia, Africa and Europe, while in North and South America, for the new cases per million, the decay in the frequency spectrum of the signal was faster than power law, except for the number of new deaths per million in the case of South America [5]. For Oceania we found the occurrence of a bifurcation in both series’ dynamics [5].

Another major finding was that all regions except Oceania showed evidence of being near a bifurcation point between a periodic window and a chaotic dynamics, also called *onset of chaos* [5]. Near the onset of chaos, chaotic attractors are characterized by low maximum Lyapunov exponents and can exhibit recurrences associated with close proximity to periodic or even quasiperiodic orbits, which become like “ghost trails” that are recurrently visited [5], leading to long evenly or unevenly spaced diagonals in recurrence matrices with 100% recurrence that only show up for a sufficiently high radius, diagonals that are intermixed with broken diagonals and isolated points which are characteristic of chaotic dynamics [5, 8].

In the case of COVID-19, the proximity of the epidemiological dynamics to a bifurcation point opens up the possibility of bifurcations occurring in the system’s dynamics with the loss of attractor stability, this occurred specifically in Oceania, as stated above, and was linked to the emergence of new variants [5].

In the present work, we expand on the work we developed in [5], combining chaos theory, machine learning and topological data analysis to address the daily hospital occupancy from COVID-19, the study compares USA with Canada, using the dataset available from Our Word in Data for the country-specific daily hospital occupancies from COVID-19 from the first available datapoint up until 2022-09-30. The methodologies applied are easily extensible to any other country. The data available for the USA includes the period from 2020-07-15 to 2022-09-30, while for Canada the period is from 2020-03-09 to 2022-09-30.

The focus on the daily hospital occupancy is relevant from a healthcare management standpoint, since the ability to use chaos theory methods to predict hospital occupancies from a pandemic, such as the SARS-CoV-2/COVID-19 pandemic, that drains/overloads hospital resources leading to deaths due to lack of resources for disease treatment, makes the prediction of hospital occupancies a critical healthcare management variable.

Secondly, from an epidemiological standpoint, hospitalizations identified as positive cases of COVID-19 are an important indicator of a double factor dynamics: viral spread among the population (the contagion) and the associated disease risk. In this way, analyzing the hospitalization dynamics from COVID-19 in terms of possible attractor properties and predictability is a pertinent and relevant research from both healthcare management and epidemiological standpoints.

Our findings show that both the USA and Canada show evidence of a stochastic chaotic dynamics characterized by power law chaos, with the USA attractor being nearer the onset of chaos than Canada.

Both countries’ attractors’ topological structure, which we characterize using topological data analysis, show strong enough recurrences to allow a high prediction performance from a forward looking adaptive A.I. system that uses topological information on these attractors and a sliding learning window, this performance does not drop significantly for multiple periods ahead prediction, indeed, the artificial agent is able to use the attractors’ recurrence structure to predict up to 6 weeks ahead with an *R*^2^ score that does not drop below 96% for the USA and 95% for Canada.

We also find that while the prediction performance is very high for both countries, showing that there is a strong deterministic pattern that can be captured for the reconstructed attractors, the USA series has a higher predictability than Canada, we explain this higher predictability through the application of both signal and topological data analysis which are convergent on the hypothesis that the USA has an attractor for the daily hospital occupancy series that is nearer the onset of chaos, which leaves a stronger periodicity topological skeleton that can be exploited by the forward looking adaptive A.I. system.

The work is divided as follows: in section 2, we review the main methods employed here, in section 3, we provide for the main results, in section 4, we provide for a final discussion on the results.

## 2. Main Methods

The main methods are divided into five main parts:

1. Signal analysis, which includes spectral analysis and signal recurrence analysis.
2. Embedding dimension estimation which will employ machine learning to choose the optimal dimension from within a dimension set.
3. Maximum Lyapunov exponent’s estimation, which will allow us to identify possible presence of chaotic dynamics.
4. Prediction performance for an adaptive A.I. system using the attractor’s recurrences for different prediction horizons, the main goal of this analysis is to evaluate the degree to which the recurrence structures contain information that can be exploited to predict the target series several periods ahead, at the same time this provides an indicator of the strength of the deterministic component.
5. Topological data analysis, which will involve *k*-nearest neighbors’ graph analysis, persistent homology analysis and recurrence analysis, which will allow us to further characterize the attractors’ topological structure.

We now describe each of these methods and their role in the work.

### 2.1. Signal Analysis Methods

The signal analysis methods that we employ are aimed at characterizing periodicities in the main series. Spectral analysis will be employed in order to find possible markers of power law dynamics, the type of memory and possible markers of high frequency periodicities that may be linked to dynamics close to the onset of chaos, this type of analysis was already employed in [5] and it proved useful for the characterization of the SARS-CoV-2’s epidemiological dynamics.

Another method is signal recurrences, in this case, we use recurrence analysis techniques applied not to the embedded series but to the original signal, this is aimed at identifying signal periodicities and possible persistent dynamics. In this case, we calculate the Euclidean distance matrix *S* for the signal *x*_*t*_ with matrix entries given by the Euclidean distance: *S*_*t,s*_ = [(*x*_*t*_ – *x*_*s*_)^2^]^1/2^ = |*x*_*t*_ – *x*_*s*_|. From this matrix we can calculate the *r*-recurrence matrix *B*_*r*_, for which an entry is equal to 0 if *S*_*t,s*_ > *r* and 0 otherwise. We calculate these *r*-recurrence matrices for different radii and, for each radii, calculate two metrics [5]: the average recurrence strength and the conditional 100% recurrence probability.

The average recurrence strength is defined as the sum of the number of points that fall within a distance no greater than the radius, in each diagonal below the main diagonal of the distance matrix *S*, divided by the total number of diagonals with recurrence below the main diagonal, this measure evaluates how strong on average the recurrence is [5, 8].

The conditional 100% recurrence probability is, in turn, defined as the probability that a randomly chosen diagonal line with recurrence has 100% recurrence, for the radius chosen [5, 8]. If all lines with recurrence had 100% recurrence, for the radius chosen, then this number would be equal to 1, the lower this metric is, that is, the closer to zero it is, the more interrupted the diagonals are, which occurs for stochastic dynamics and also for deterministic chaotic dynamics, as discussed in [5, 8].

These two metrics allow us to further characterize a signal’s periodicities and how strong recurrences are, becoming a signal topological analysis tool complementary to the spectral analysis. This analysis also allows us to select possible radii for the machine learning component.

### 2.2. Delay Embedding and Largest Lyapunov Exponent Estimation

Delay embedding involves embedding a time series in a point cloud in a multidimensional Euclidean phase space, the resulting embedded trajectory can be research upon, including the possibility of presence of a dynamical attractor [5, 9, 10]. The use of delay embedding as an attractor reconstruction method working from a time series *x*_*t*_ may be able to recover the main properties of the attractor, a point that comes from Takens’ theorem [10], delay embedding can also be employed as a feature space for time series prediction [5, 9]. Using an appropriate time delay *σ* and embedding dimension *d* in Euclidean space, the delay embedding involves building a sequence of *d*-dimensional tuples from the time series:

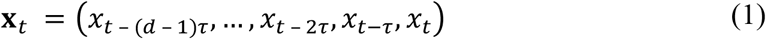

The resulting trajectory of the phase point **x**_*t*_ may contain topological regularities, using the Euclidean topology, that can be exploited by machine learning algorithms to predict the target series, an approach that was employed in [5] to predict with success the COVID-19’s new cases per million and new deaths per million series, both in cases where there was evidence of an attractor and in the Oceania case, for which a bifurcation occurred [5]. For a delay embedding, the delay choice should be linked to the memory and characteristics of the dynamics. As argued in [5], in epidemiological contexts, we can use incubation period data and possible quarantine windows, if a quarantine period is established or recommended by healthcare authorities and implemented by governments, in this case, the first day after a recommended quarantine period allows for the embedding to account for quarantine effects, this point was addressed and argued in [5] in regards to SARS-CoV-2, where the World Health Organization (WHO) recommended quarantine period is 14, leading to a 15 day period lag, so that the 15 day lag is the number of days between the start and the end of the recommended 14 day quarantine period [5].

Now, to obtain a phase space embedding that provides predictable features for a target series we can use a machine learning method to select the embedding dimension, this method was employed in [5] to deal with the case of Oceania, where a bifurcation occurred, for which no stable attractor assumptions associated with traditional phase space embedding selection methods apply [9]. In this case, in [5], we calibrated the embedding dimension from a set of alternative dimensions to the one that gave the best result in the prediction of the target series.

This method can also be employed when the dynamics is in an attractor. In this case, using a nearest neighbors’ machine learning algorithm, either a radius learner or a *k*-nearest neighbors’ learner, we can build a prospective prediction A.I. system that uses the topological regularities in the embedded trajectory to predict the target series. We can then select, from a set of alternative embedding dimensions, the one that leads to the best prediction performance. In this way, we make sure that we have an embedding that captures the most of the predictable topological structure of an attractor, when an attractor is present, allowing us to further study the topological properties of the attractor using topological data analysis methods, a point that, as shown in [5], also applies to the cases where dynamical changes are present, for which the embedding that leads to the highest results in prediction can be used as a base embedding to study the topological changes that occurred.

The adaptive A.I. technology that we use is based on a sliding window prospective machine learning model which, as stated, has been successfully applied in epidemiological prediction including SARS-CoV-2 [5, 11]. Since we will be using topological data analysis based on the Euclidean distance matrix, we use a Euclidean radius learner for the A.I.’s adaptive processing, employing Python’s machine learning’s library’s scikit-learn’s radius learner.

In the case of the series studied in the present article, there are no identifiable bifurcations, so this method is able to find an embedding dimension, from a set of dimensions, that provides for the best prediction results, so that, when we apply the topological data analysis, we are applying it to the embedding that leads to the highest exploitable topological information for the target series prediction, and, in this way, we are not only assured that we have the embedding dimension that leads to the highest amount of topological information from the studied set of alternative embeddings, we can also link the topological analysis directly to the predictability of the target series, which from an epidemiological and healthcare management standpoint is a key advantage.

Following the approach addressed above and also employed in [5], we perform different embeddings and, for each embedding dimension *d*, we use an Euclidean radius-based learner with a sliding learning window of size *w*, to perform the single period prediction:

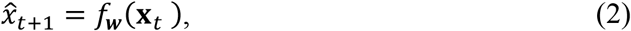

using as training data the sliding window feature set {x_*t*−*w*−1_, …, x_*t*−1_} and sliding window target {*x*_*t*−*w*_, …, *x*_*t*_}. We employ sciki-learn’s radius neighbor regressor using the Euclidean metric, in order to test within a set of dimensions *d*_0_, *d*_1_,…,*d*_*N*_ which embedded dynamics leads to the best prediction performance of the target series. In this case, we use the *R*^2^ score as a metric for selecting the dimension, this dimension leads to the embedding for which a radius learner is able to extract the most information from the topological structure of the embedded trajectory to predict the target series, having obtained such an embedding, we apply Rosenstein *et al*.’s method for the estimation of the largest Lyapunov exponent, a positive largest Lyapunov exponent being a marker of chaotic dynamics [5].

A positive largest Lyapunov exponent indicates the presence of sensitive dependence upon initial conditions, in the case of stochastic chaos with a low dimensional attractor, the higher the value of this exponent is, the more sensitive is the dynamics to small noise fluctuations and the lower is the prediction horizon [5,6]. In the case of SARS-CoV-2, in [5], we found that the new cases per million series and the new deaths per million series, for the regional data where an attractor emerged, were characterized by low values of the largest Lyapunov exponents, consistent with a dynamics close to the *onset of chaos*.

Low values of the largest Lyapunov exponents, coupled with color chaos signatures with long range persistent dynamics leads to a strong long-range predictability that can be exploited by forward looking adaptive A.I. algorithms, such as the one we use here, taking advantage of the recurrence structures associated with these attractors [5]. Furthermore, color chaos dynamics close to bifurcation points near periodic windows (*onset of chaos*) have recurrence signatures associated with a close proximity of cycles that make these dynamics more predictable in the long range, which leads us to the next point which is the test of the prediction performance on multiple prediction horizons, an analysis which is of relevance for healthcare management since it addresses how A.I. solutions can be deployed for early warning systems on healthcare resource usage, also, from an epidemiological standpoint, it provides for relevant insights since a high long term predictability, in the case of hospitalization data, provides information on the virus’ infectiousness patterns.

### 2.3. Prediction Performance for Multiple Prediction Horizons

We begin by analyzing the prediction performance of the forward looking adaptive A.I. system, reviewed above, on a one-day ahead prediction horizon, reporting the following as main metrics [5]:

1. The linear correlation between the A.I. predictions and the target signal, which allows us to evaluate how much the A.I. predictions match the fluctuations in the target variable, a score that should be positive and high for a well performing prediction technology; The root mean squared error divided by the total data amplitude, which provides for a relative error measure and thus gives a relative scale on error; The explained variance score, which is one of the main metrics providing for the degree to which the variability in the target is explained by the A.I.’s predictions; The coefficient of determination (*R*^2^) score, which is similar to the explained variance score, but accounts for systematic offsets in prediction, in this way, being preferred to the explained variance score.
2. The above metrics, when calculated for an adaptive A.I. system, with a sliding window radius learning unit, can be used to characterize the degree to which the topological structure of the attractor, in terms of recurrences, can be exploited to predict the target variable, providing for a more detailed picture of the prediction performance, in this way, it can also be employed to evaluate the level of regularities in the recurrence structure of the reconstructed attractor that contains information on the next period value of the target series.
3. Going beyond the one-day ahead prediction horizon, we calculate the *R*^2^ score on multiple prediction horizons where the A.I., instead of being tasked to learn to predict the target one day ahead, is tasked with predicting the target several days ahead, using the embedded trajectory [5].
4. From an healthcare standpoint, when dealing with hospital occupancy from COVID-19 as target of interest, a multiple periods ahead prediction is a key analysis, since, if the performance is good, we can use the reconstructed attractor to predict hospitalizations several days/weeks ahead offering for a foresight that can be used by hospital management for planning, it also offers a country’s healthcare authorities a foresight into possible hospital resources’ pressure that can guide healthcare planning and response, finally, from an epidemiological standpoint, it offers insight into how the virus is behaving in terms of morbidity, offering insights into patterns associated with topological regularities that occur in the dynamics of a possible chaotic attractor associated with the hospitalizations themselves.

### 2.4. Topological Data Analysis

The topological data analysis methods complement the previous analyses. The first analysis that we perform is based on *k*-nearest neighbors, in this case, we test, first, a similar A.I. for the one-day ahead prediction to that of the previous subsection but replacing the radius learner by a *k*-nearest neighbors’ learner, evaluating, for different values of *k* and a sliding learning window, the value of the *R*^2^ score, this serves a double purpose, one is to assess the degree to which the *k*-nearest neighbors of an embedded trajectory contain information that may help such an adaptive forward looking A.I. system to predict the future value of the target series, the second purpose is to select the best value of *k* to analyze the *k*-nearest neighbors’ graph **N** for the reconstructed attractor, this method was successfully employed in [5] to analyze the reconstructed attractors for the regional series of the number of new positive cases per million and the number of new deaths per million from COVID-19.

The *k*-nearest neighbors’ graph **N** is an undirected graph with the vertices corresponding to each phase point and the edges corresponding to the *k* nearest neighbors. From the graph **N**, one can extract the set of degree values *J*, and the degree distribution, calculating the relative frequencies *p*_*j*_ associated with each degree value *j* ∈ *J*, from this distribution, the degree (relative) entropy can be calculated as [5]:

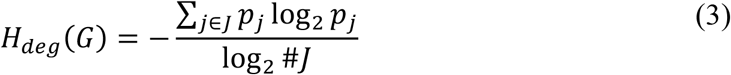

This relative entropy measure has a value between 0 and 1, in the special case of a graph where the relative frequencies associated with each degree coincide *p*_*j*_ = 1/#*J, H*_*deg*_(*G*) = 1, which is the maximum entropy value, in the case of a graph where each node has the same degree we get *H*_*deg*_(*G*) = 0, which is the lowest entropy value. The closer to 1 this degree entropy measure is, the closer the graph is to a maximum degree entropy.

The Kolmogorov-Sinai (K-S) entropy is the second graph entropy measure that we calculate for the *k*-nearest neighbors’ graph, this entropy measure is an information measure for a Markov process with a transition matrix extracted from the graph. For an unweighted graph, which is our case, this entropy coincides with the logarithm of the dominant eigenvalue of the transition matrix *μ*_+_, expressing it in bits, leads to the following information measure [5,12]:

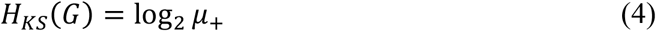

The last topological data analysis method that we employ is persistent homology [5, 13, 14], worked from the Euclidean distance matrix *S*, calculated on the embedded series, which includes all the Euclidean distances between the embedded points in *d*-dimensional Euclidean space, allowing one to find relevant topological features in the embedded trajectory and analyze how the homology changes over a Vietoris-Rips filtration calculated on the embedded series [5]. In this case we look at the 0, 1 and 2-homology classes. The 0-homology class (H0) corresponds to components connected by a line, therefore having a zero dimensional boundary, a 1-homology class (H1) corresponds to a loop, finally the 2-homology class (H2), correspond to voids, that is, simplexes with faces but no interior [5, 13, 14].

Persistent homology, can be used to count the number of structures in each simplicial complex in a Vietoris-Rips filtration for each homology dimension, including the birth and death of homology classes as the radius is increased. The homology classes’ birth and death can be calculated in the following way, given a filtration of simplicial complexes 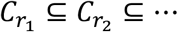, we get a sequence of maps for the homology dimension *s*, 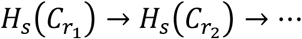, a homology class is born at *n* if it is in 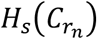 but not in the image of the map 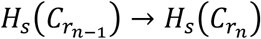 and dies at *m* if it is in 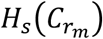 but not in 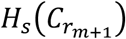 [13].

Persistent structures (*long lived classes*) are large scale structures in the embedded trajectory, if that trajectory is in an attractor, they correspond to large scale structures of the attractor, structures with lower persistence may be indicative of the presence of noise or of a stronger chaotic dynamics [5]. A persistence diagram *D* can be calculated for different homology dimensions with each point for each dimension giving the lifetime for a structure, in this case, following the methodology employed in [5], we define *D*_*S*_ as the persistence sub-diagram for the homology dimension *s*, each point in the sub-diagram corresponds to an ordered pair of birth and death times in the filtration.

The lifetime or persistence of a class at dimension *s*, thus, corresponds to the difference between the death and birth times, therefore, given a dimension *s* and the ordered pairs of the sub-diagram (*n*_*B*_, *n*_*D*_) ∈ *D*_*S*_, where *n*_*B*_ is the “filtration birth time” and *n*_*D*_ is the “filtration death time”, with the death happening after birth, we can calculate the persistence metric as [5,13]:

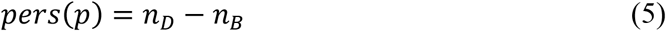

Structures that live through the full filtration have *n*_*D*_ = ∞ and, therefore, we get *perS*(*p*) = ∞. Now, to better characterize the attractor’s topology we apply the same methodology applied in [5] and calculate the following metrics for each homology dimension (that is, for each sub-diagram *D*_*S*_):

1. The number of classes with *perS*(*p*) < ∞, which allows us to identify which dimension is predominant in terms of number of homology classes with lifetimes shorter than ∞;
2. The number of classes with *perS*(*p*) = ∞, which allows us to identify the homology dimensions that have structures that persist throughout the whole filtration, constituting very large scale structures;
3. The maximum persistence which allows us to characterize which dimensions have the most persistent structures;
4. The mean persistence: this metric allows us to characterize each homology dimension in terms of its mean persistence.

With these metrics calculated for the different sub-diagrams we get a picture of an attractor’s structure at multiple dimensions and the dominant features, an approach that was applied in [5].

A final analysis that we perform is again to calculate the average recurrence strength and the conditional probability of 100% recurrence but on the Euclidean distance matrix calculated for the embedded series in *d*-dimensional Euclidean space, the same matrix used for the persistent homology analysis.

## 3. Results

In figure 1 we show the USA and Canada’s hospital occupancy numbers from COVID-19, for the periods from 2020-07-15 to 2022-09-30 and from 2020-03-09 to 2022-09-30, respectively.

**Figure 1:**
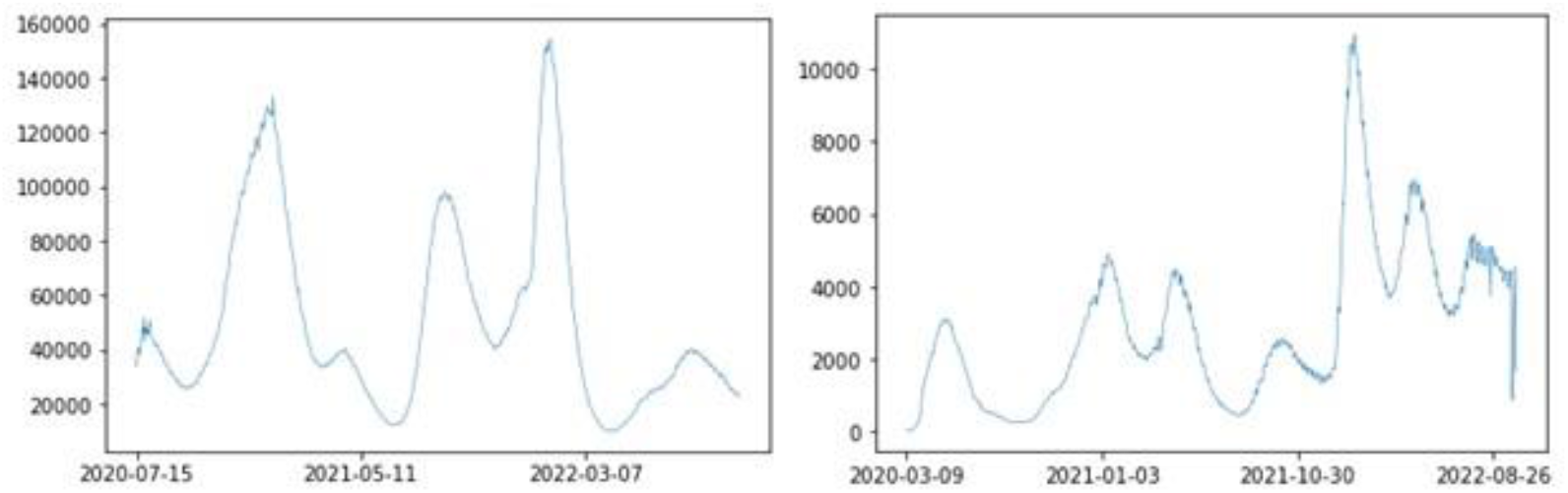
Daily hospital occupancy from COVID-19 in USA (left), from 2020-07-15 to 2022-09-30, and Canada (right), from 2020-03-09 to 2022-09-30.

In figure 2, we show the power spectrum for both countries, in both cases we find the presence of a power law decay in the spectrum with an estimated exponent of 4.469975 for the USA, with an associated *p-value* of 3.573819e-29 and an *R*^2^ of 0.895767, which implies a strongly persistent spectrum, for Canada the spectrum is also strongly persistent but slightly less than the USA, with an estimated exponent of 3.866758, with an associated *p-value* of 4.076911e-24 and *R*^2^ of 0.842180. There is a slight rise with a peak at the high frequency window for both countries, which is indicative of a high frequency signal, this can happen in chaotic attractors near the onset of chaos, where the dynamics is near a periodic window, which can lead to high frequency markers [5], such a dynamics was found to occur for North America’s COVID-19 new daily positive cases per million and new daily deaths per million in [5], which may also explain the daily hospital occupancies from COVID-19’s high frequency markers.

**Figure 2:**
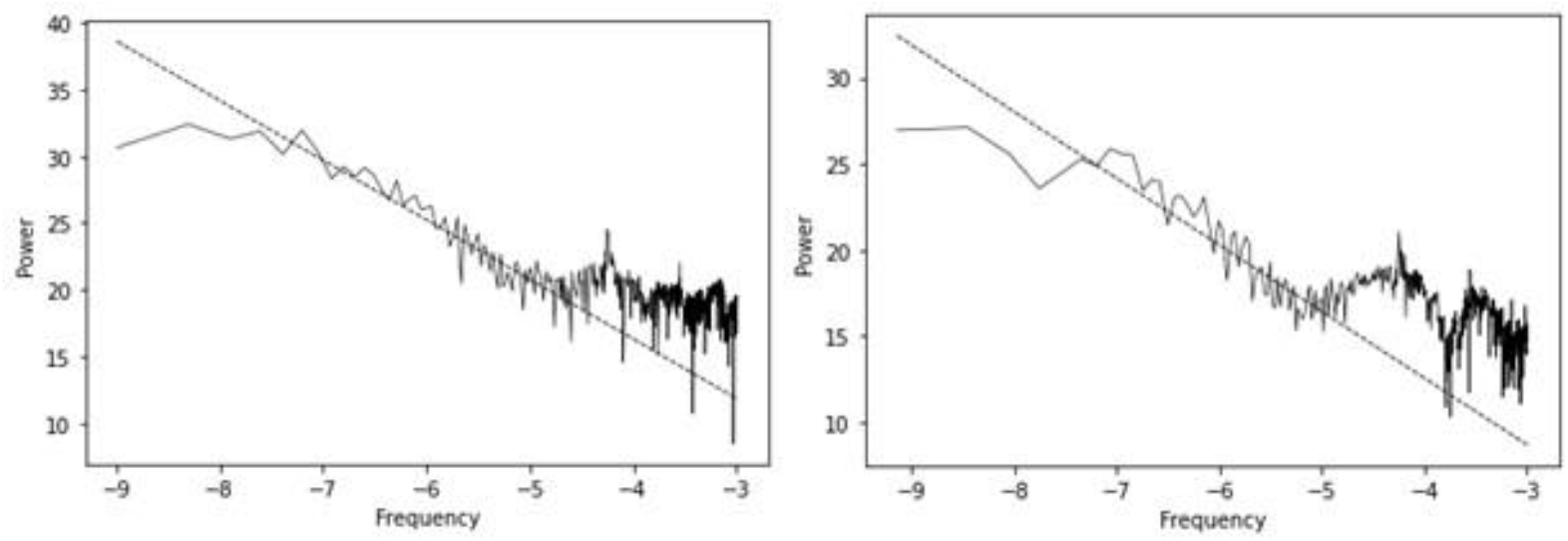
Power spectra for the daily hospital occupancy from COVID-19 series for the USA (left) and Canada (right).

The strong persistence of the signal, and the high frequency signatures indicate that there may be a recurrence structure that is strong enough for an adaptive A.I. system to predict the series, using an appropriate phase space embedding.

In table 1, we show the average recurrence strengths for different radii in units of standard deviation. We find that the USA has, predominantly, a higher average recurrence strength than Canada, which is also consistent with the higher persistence found in the power law decay in the spectral analysis. With increasing radius, the two countries’ average recurrence strengths converge. The conditional 100% recurrence probability for the USA is also higher than that of Canada, remaining so with increasing radii, as shown in table 2, which reinforces the above results.

**Table 1:** Average recurrence strengths with increasing radii in units of standard deviation for the original series.

| Radius | USA | Canada |
| --- | --- | --- |
| 1 | 0.653486 | 0.470163 |
| 1.5 | 0.754103 | 0.656440 |
| 2 | 0.840627 | 0.818387 |
| 2.5 | 0.909904 | 0.921389 |
| 3 | 0.961158 | 0.956449 |
| 3.5 | 0.989799 | 0.973040 |
| 4 | 0.998442 | 0.985131 |

**Table 2:** Conditional 100% recurrence probability calculated for different radii with increasing radius in units of standard deviation.

| Radius | USA | Canada |
| --- | --- | --- |
| 1 | 0.151177 | 0.001070 |
| 1.5 | 0.166047 | 0.002139 |
| 2 | 0.187113 | 0.035294 |
| 2.5 | 0.288724 | 0.165775 |
| 3 | 0.394052 | 0.223529 |
| 3.5 | 0.583643 | 0.344385 |
| 4 | 0.863693 | 0.473797 |

In this last case, it is noticeable that the USA has a significantly higher value than Canada which is indicative of a closer proximity to a periodic or quasiperiodic skeleton associated with a periodic or quasiperiodic window, that leaves something like a *ghost trail* in the chaotic dynamics, another indicator favorable to the hypothesis that the USA dynamics may be closer to the onset of chaos.

Considering the predictability of the dynamics, using delay embedding, with a 15 period lag and a 7-days sliding learning window, as shown in table 3, for embeddings varying from 2 to 10 and a radius learner, we find that all the embeddings lead to a high value of *R*^2^ (higher than 90%), which means that the recurrences contain sufficient information for a high predictability of the hospital occupancies from COVID-19, using a radius learner, in all applications of machine learning, in the present work, we use a brute force algorithm and an Euclidean metric.

**Table 3:** *R*^2^ scores for the radius adaptive learner, using a 7-days sliding learning window and a radius of 4 *s.d*., with increasing embedding dimensions.

| dE | USA | Canada |
| --- | --- | --- |
| **2** | 0.964440 | 0.952165 |
| **3** | 0.964498 | 0.952816 |
| **4** | 0.964381 | 0.953132 |
| **5** | 0.964055 | 0.953183 |
| **6** | 0.963702 | **0.953381** |
| **7** | 0.963603 | 0.953370 |
| **8** | 0.963853 | 0.952805 |
| **9** | **0.965104** | 0.951742 |
| **10** | 0.965092 | 0.950427 |

**Table 4:** Largest Lyapunov exponents estimated for the USA and Canada’s embedded series.

|  | **dE** | **L1** |
| --- | --- | --- |
| **USA** | 9 | 0.007729 |
| **Canada** | 6 | 0.014471 |

In the case of the USA, we find that the highest *R*^2^ (around 96.51%) holds for a nine dimensional embedding, while for Canada the highest *R*^2^ (around 95.34%) holds for a six dimensional embedding. Using these embedding dimensions, 9 for the USA and 6 for Canada, we find that the corresponding estimated largest Lyapunov exponents are both positive, which is a signature of chaos associated with the daily number of hospital occupancies from COVID-19, in this case, given the spectral signatures we find that we may be dealing with color chaos, however, for the hospital occupancy series, while both countries show evidence of chaos, the estimated value of the largest Lyapunov exponent is smaller for the USA than for Canada, which means that the USA dynamics may be closer to the *onset of chaos* than that of Canada.

In this way, the evidence is favorable, for both countries, to a hypothesis of a chaotic dynamics with a high degree of predictability by an adaptive A.I. system that uses the embedded series’ recurrences to predict the daily number of hospital occupancy from COVID-19, the associated dynamics is consistent with a form of stochastic chaos characterized by a power law decay in the power spectrum (color chaos), with high frequency periodic signatures and low values of Lyapunov exponents consistent with the chaotic dynamics being close to the onset of chaos, with the USA being characterized by a higher dimensional structure than that of Canada, stronger recurrences, higher predictability and a Lyapunov exponent that is closer to zero, indicating that the USA’s possible chaotic attractor may be closer to the onset of chaos.

Considering now, the change in predictability with the sliding learning window, we find that the *R*^2^ values for the radius learner decrease with the window size, in this way the sliding 7-day learning window (one week) for the adaptive A.I. system leads to the best performance (table 5), which means that the one week learning window should be preferred. Similar results were obtain in [5], with the one week learning window also leading to a better performance for COVID-19’s new cases and new deaths per million.

**Table 5:** *R*^2^ scores for the radius adaptive learner, using a radius of 4 *s.d*., with increasing window sizes.

| Window | USA | Canada |
| --- | --- | --- |
| **7** | **0.965104** | **0.953381** |
| **8** | 0.958129 | 0.945731 |
| **9** | 0.950557 | 0.937400 |
| **10** | 0.942484 | 0.928642 |
| **11** | 0.933947 | 0.919588 |
| **12** | 0.924948 | 0.910263 |
| **13** | 0.915447 | 0.900572 |
| **14** | 0.905456 | 0.890304 |
| **15** | 0.894992 | 0.879485 |

Going beyond the one-day ahead prediction, we find that the Euclidean recurrence structure contains sufficient information to allow the adaptive A.I., equipped with a radius neighbor machine learning module, to predict the future hospital occupancies from COVID-19 for longer horizons, as shown in figure 3.

**Figure 3:**
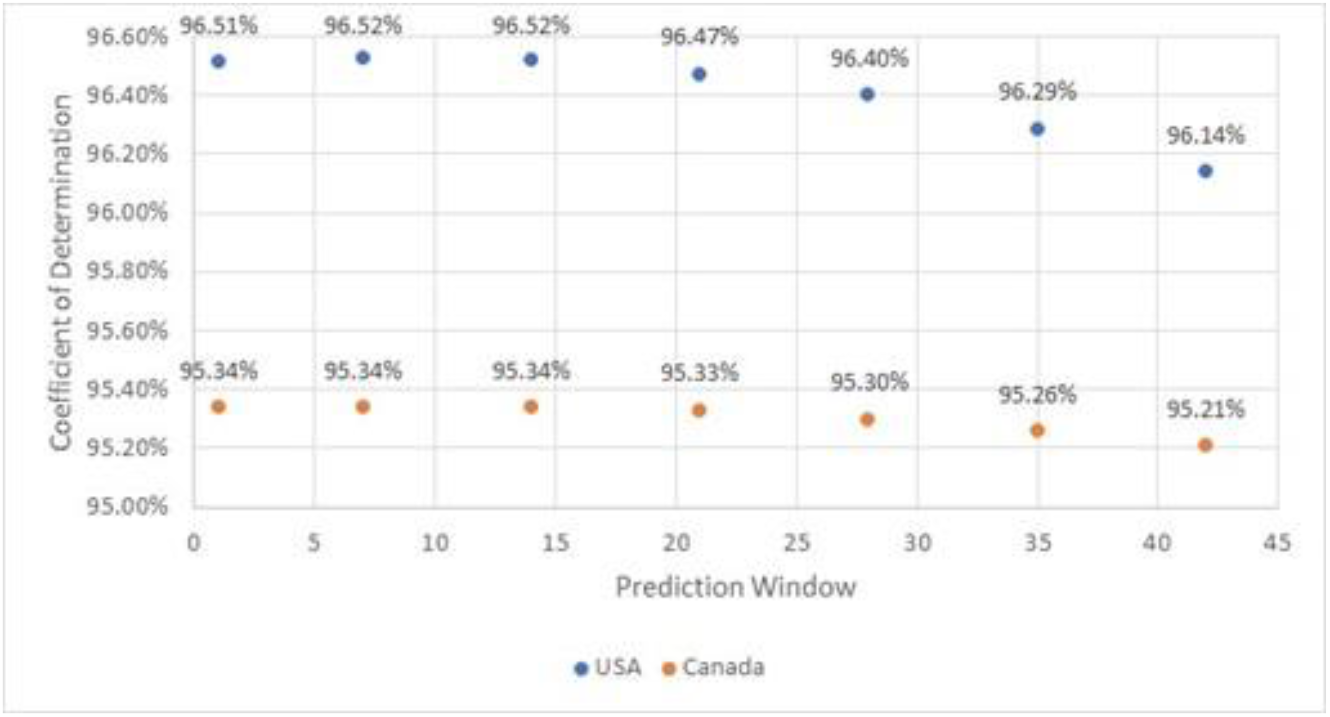
*R*^2^ scores for prediction windows 1, 7, 14, 21, 28, 35 and 42 days ahead, using a 7-days sliding learning window.

Indeed, counting the weeks in terms of 7-day periods, we find that up to two weeks ahead (14 days) the adaptive A.I.’s prediction performance, measured in terms of the *R*^2^, does not drop with respect to the single day prediction horizon, only after 14 days do we see a break in prediction performance, however, that break is not significant, indeed, up to 6 weeks ahead (42 days) we find that the prediction performance for the USA hospital occupancy from COVID-19 does not drop below 96%, and for Canada it does not drop below 95%. These values show that the recurrence structures for the reconstructed attractors allows for the development of of A.I. systems that exploit them in a way that can be deployed by healthcare management authorities to predict the future hospital occupancies and take appropriate measures. Again, as in the previous analysis, the USA series has a higher predictability than Canada.

Considering, now, the A.I. equipped with the *k*-nearest neighbors’ algorithm, we find that the performance is higher than that of the radius neighbors’ learner and that it decreases with increasing *k* (table 6), with the best performance obtained for *k* = 2 nearest neighbors, we will thus use this value in the *k*-nearest neighbors’ topological analysis. Once more, the performance for the USA is higher than for Canada.

**Table 6:** *R*^2^ scores of the *k*-NN adaptive A.I. for the USA and Canada’s daily hospital occupancies from COVID-19 embedded series, using a 7-days sliding learning window.

| $k$ | USA | Canada |
| --- | --- | --- |
| **2** | **0.989918** | **0.978189** |
| **3** | 0.986178 | 0.974072 |
| **4** | 0.981870 | 0.969883 |
| **5** | 0.976961 | 0.965221 |
| **6** | 0.971384 | 0.960017 |

The higher prediction performance for the USA than for Canada can be further accounted for by employing the *k*-nearest neighbors’ topological analysis. In figure 4, we show the *k*-nearest neighbors’ graphs, for *k* = 2, for both countries and the respective degree distribution. We find some differences between the two graphs, while, as shown in table 7, both graphs exhibit low degree entropy values, the graph for the USA has a lower degree entropy than the graph for Canada, the same is true of the *K-S* entropy which is lower for the USA than for Canada, furthermore, the *k*-nearest neighbors’ graph for the USA is not scale free (power law scaling), while Canada’s graph shows a region of power law scaling in the degree distribution, which implies the possible presence of a scale free graph in the neighborhood structure.

**Table 7:** Main entropy values for figure 4’s *k-*NN graphs.

|  | USA | Canada |
| --- | --- | --- |
| **Degree Distribution Entropy** | 0.047259 | 0.123603 |
| **K-S Entropy** | 1.479485 | 1.781877 |

**Figure 4:**
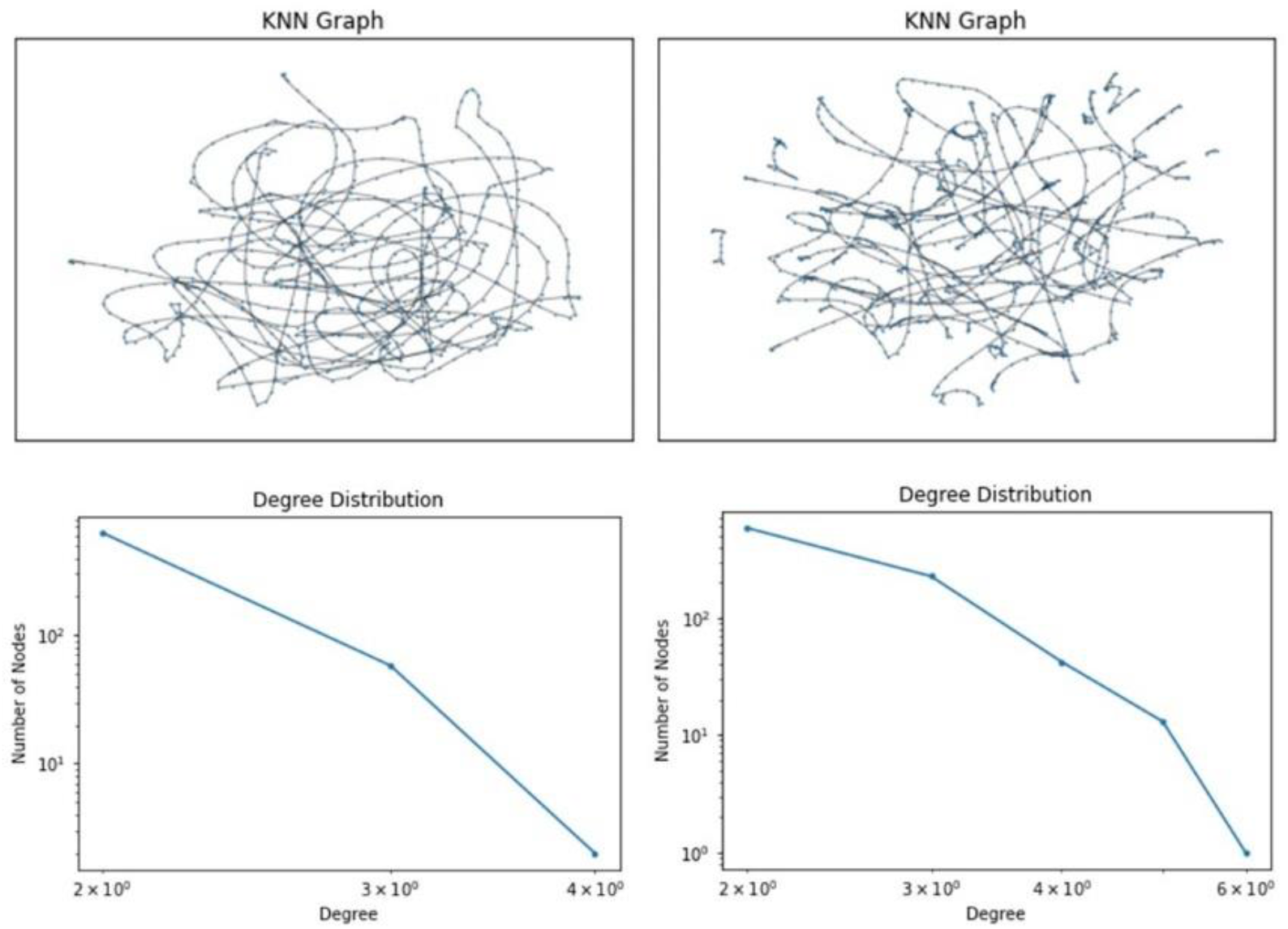
*k*-nearest neighbors graphs and respective degree distribution for the USA (left) and Canada (right).

The lower entropy values of the graph structure and the non-power law decay for the USA’s degree values may be indicative of a lower complexity of the USA’s nearest neighbors structure and that the USA’s attractor is closer to a bifurcation point from a periodic window to a chaotic dynamics, which is consistent with the previous result from the largest Lyapunov exponent that shows that the USA has an estimated exponent closer to zero than Canada. In this way, the evidence is favorable to the USA’s daily hospital occupancy from COVID-19’s attractor being closer to the onset of chaos than Canada’s. Thus, even though the evidence is favorable for the USA’s attractor having a higher dimensionality, the evidence also supports the hypothesis that this attractor is closer to the onset of chaos.

Considering now the persistent homology analysis, we show in figure 5 the distance matrices and respective persistence homology diagrams, the distance matrices are in color code, where the lighter colors correspond to the smaller distances and the darker colors to larger distances.

**Figure 5:**
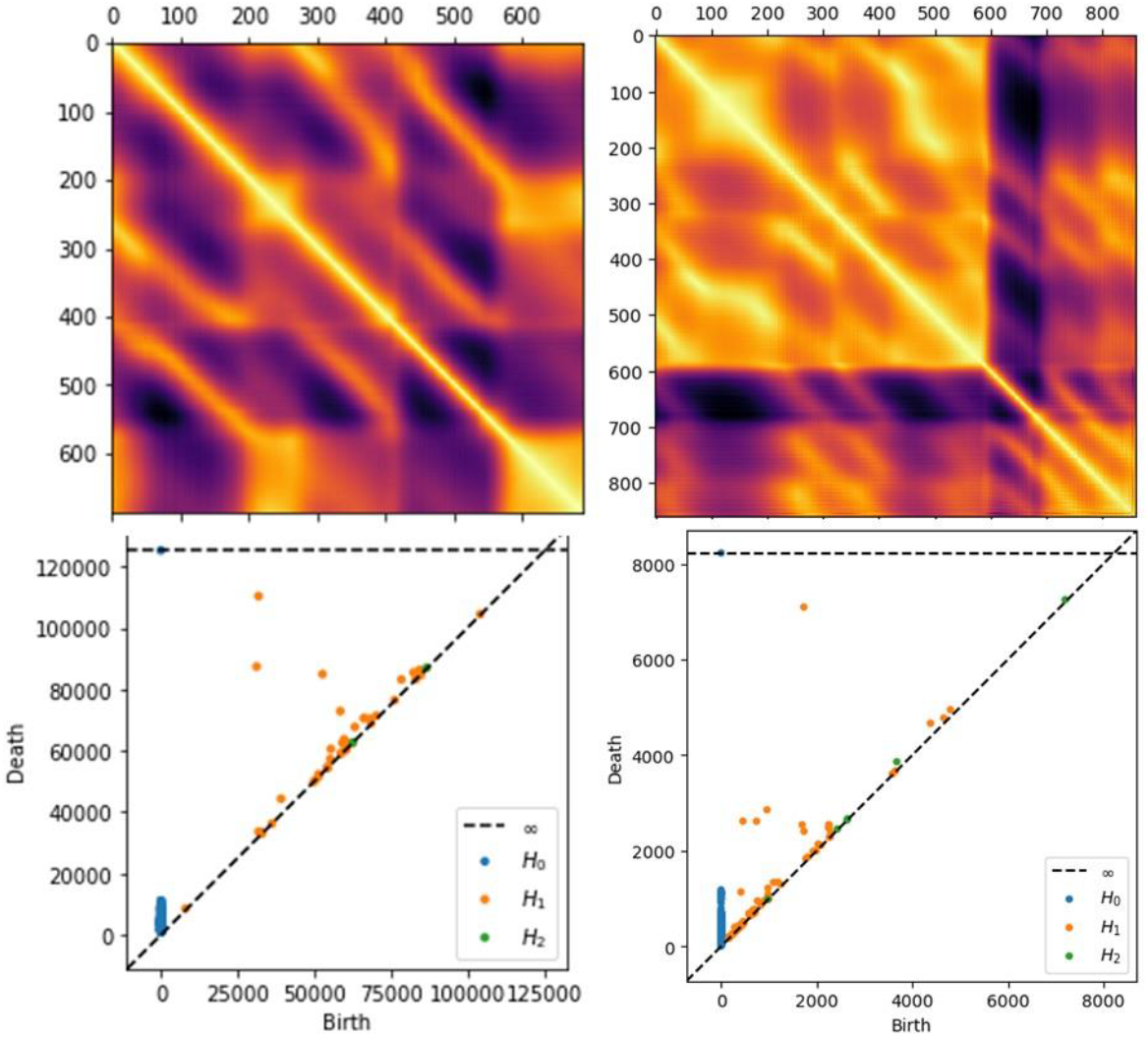
Distance matrices (top) and respective persistence diagrams (bottom) obtained from the embedded series for the USA (left) and Canada (right).

There is an immediately visible difference between the two countries, the homology dimensions 1 and 2 are born much sooner in the filtration for Canada than for the USA, however, the similarities and differences between the two countries, with respect to the persistence diagrams, become more visible when we consider the persistence metrics extracted from the respective diagrams, as shown in tables 8 and 9. With respect to the number of classes, we already find a difference between the two countries, indeed, while both countries have a predominance of classes of homology dimension 0, followed by homology dimension 1 and, finally 2, as the more residual dimension, Canada’s attractor has a higher number of classes than the USA attractor, also, for homology dimension 2, Canada’s attractor has 6 classes while the USA only has 2 classes. Both countries have one infinity class that holds for homology dimension 0, and the maximum persistence, that is not an infinity class, is obtained in both countries for homology dimension 1, which indicates that there is a presence of loops in the attractor’s larger scale topological structure.

**Table 8:** Persistence metrics for the USA attractor.

|  | **H0** | **H1** | **H2** |
| --- | --- | --- | --- |
| **Number of Classes** | 688 | 44 | 2 |
| **Maximum Persistence** | 11,115.62 | 78,518.19 | 294.5 |
| **Mean Persistence** | 4,356.412 | 5,342.618 | 161.621 |

**Table 9:** Persistence metrics for the Canada attractor.

|  | **H0** | **H1** | **H2** |
| --- | --- | --- | --- |
| **Number of Classes** | 861 | 80 | 6 |
| **Maximum Persistence** | 1,182.684 | 5,365.721 | 186.550 |
| **Mean Persistence** | 268.919 | 225.058 | 48.509 |

At the mean persistence level, we find the biggest difference between the two attractors, indeed, the USA has homology dimension 1 as the predominant in terms of persistence structures, which means that loops are larger scale structures for its attractor, indicating, again, a possible closer proximity to a periodic window.

For Canada, homology dimensions 0 and 1 are close to each other in terms of mean persistence, but homology dimension 0 has a higher value in terms of mean persistence.

To complement this analysis, considering a radius of 4 s.d., we find that both countries have a high recurrence probability (table 10), however, the embedded series for Canada has a higher recurrence probability than for the USA, it also has a higher average recurrence strength, however, the USA has a higher probability of finding 100% recurrence diagonal lines in lines with recurrence (15.5146% probability for the USA, against 9.9644% probability for Canada), which again reinforces the hypothesis of a closer proximity of the USA to a periodic window, leaving a stronger marker in the probability of finding 100% recurrence lines, conditional on these lines being lines with recurrence.

**Table 10:**
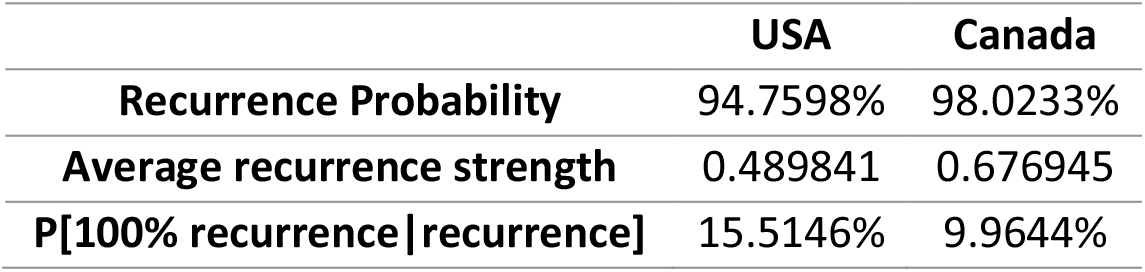
Recurrence metrics using a radius of 4 s.d. for the USA and Canada’s embedded series.

## 4. Conclusions

We applied chaos theory and topological data analysis methods combining A.I. with *k*-nearest neighbors, persistent homology and recurrence analysis to USA and Canada’s daily hospital occupancies from COVID-19. The results show that there is evidence of the emergence of a low dimensional attractor for both countries with the best fit in A.I. target prediction being obtained, in the case of the USA, for a 9 dimensional embedding and in the case of Canada for a 6 dimensional embedding, out of a tested range of dimensions up to 10.

In both cases, we find that the largest Lyapunov exponent estimated for the reconstructed attractor is positive, which is an indicator of a chaotic dynamics at the level of the daily hospital occupancies from COVID-19. Furthermore, despite the lower dimensionality, Canada’s attractor exhibits a higher Lyapunov exponent and is less predictable than the USA attractor, even though both attractors’ recurrences can be exploited for a long-range predictability by an adaptive forward looking A.I., which means that healthcare authorities can implement A.I. solutions using delay embedding to predict hospital occupancies from COVID-19 and plan for healthcare responses. Of notice, we found that adaptive A.I. systems that exploit the reconstructed attractor recurrences employing epochal learning via a sliding learning window can predict the hospitalization numbers for both countries with more than 95% *R*^2^ score up to 42 days ahead, which means that the attractor reconstruction coupled with such machine learning solutions allow the implementation of early warning systems for hospital resource utilization associated with hospital occupancies from COVID-19.

We traced down this predictability to the respective attractors’ topological structures. Our analysis supports the hypothesis of a form of noisy power law (color) chaos in both the USA and Canada’s daily hospital occupancies from COVID-19, but with the USA being closer to the onset of chaos than Canada, this shows up in the topological data analysis, spectral analysis, Lyapunov exponents, recurrence metrics (at the level of the conditional 100% recurrence probability) and persistent homology analysis.

Canada’s attractor is more chaotic than the USA, so that, while still having strong recurrences that can be exploited for prediction, the adaptive A.I. system shows consistently lower performance, the Lyapunov exponent is higher, the power law decay in the frequency spectrum of the signal is faster and the *k*-nearest neighbors graph shows a scale free degree distribution, which the USA attractor does not.

The current findings are convergent with our previous work on the regional data for the COVID-19 number of new cases per million and new deaths per million, which showed the presence in North America of chaotic attractor structures close to the onset of chaos, even though the decay in the frequency spectra was faster than the power law.

The methods we employed here are scalable for other countries and are also adaptable in setting embedding parameters for cases where bifurcations take place, as shown in [5] for the case of Oceania’s new cases per million and new deaths per million series. The methods can also be employed for other diseases.

## Data Availability

All data produced are available online at:
https://ourworldindata.org/covid-hospitalizations
The download date set the last datapoint at 2022-09-30.

